# Dengue virus outbreak in Cochabamba, Bolivia

**DOI:** 10.1101/2025.05.19.25327817

**Authors:** Paola Mariela Saba Villarroel, Selin Sen, Raphaëlle Klitting, Laura Pezzi, Geraldine Piorkowski, Norma Villavicencio Siles, Sineewanlaya Wichit

## Abstract

With *Aedes aegypti* expanding to higher altitudes, dengue virus (DENV) cases soared to record levels in 2024 in Cochabamba, Bolivia (2,558 meters above sea level). Using genome sequencing, we identified at least two distinct clades of DENV-2 genotype II circulating during the outbreak.

## Introduction

Dengue virus (DENV), a member of the *Orthoflavivirus* genus (*Flaviviridae* family), comprises four serotypes (DENV-1 to DENV-4) (1), each further subdivided into several genotypes. Among them, DENV-2 includes six genotypes, with genotype II (Cosmopolitan) being the most widespread and genetically diverse (1,2). DENV is endemic across most tropical and subtropical regions of the world, where its primary mosquito vector, *Aedes (Ae*.*) aegypti*, is present. In the Americas, dengue cases have surged dramatically in recent years, increasing by 361% in 2024 compared to the five-year average (3), with more than 12 million suspected cases reported in the region that year (4).

In Bolivia, the virus has been circulating since 1931, with seroprevalence studies revealing high infection rates in the tropical and subtropical lowlands by 2017, including Santa Cruz (93.5%) and Beni (90.0%), and lower rates in highland regions such as Cochabamba (10.4%) and La Paz (11.8%) (5). In recent years, *Ae. aegypti* has expanded its geographic range to nearly two-thirds of the country, reaching altitudes of up to 2,700 meters in the Andes (6). Along with this expansion, a significant rise in annual dengue cases has been observed in areas that were previously less affected. Notably, in Cochabamba, confirmed cases rose from fewer than 130 before 2018 to 1,400 in 2019. In 2024, Cochabamba experienced an unprecedented outbreak, with 13,022 suspected and 8,087 confirmed cases reported by December, accounting for 63.2% of Bolivia’s total confirmed infections (7).

## The study

Cochabamba Department, Bolivia’s third most populous region, has approximately 2 million inhabitants. Its capital, Cochabamba city, located in Cercado Province, is situated in a valley within the Andes mountains at an altitude of 2,558 meters and is the most densely populated area of the department, home to 661,484 residents as of 2024. Cochabamba city experiences a semi-arid climate (Köppen: BSk), bordering on a subtropical highland climate (Köppen: Cwb), characterized by a dry season from May to October and a wet season from November to March. Another important province, Quillacollo, located at 2,422 meters above sea level, has been undergoing rapid urban and demographic growth (8–10).

Between January 2 and July 15, 2024, a total of 9,576 suspected DENV cases were analyzed at the Departmental Health Service (SEDES), Cochabamba, using DENV_NS1 ELISA, MAC-ELISA IgM, and/or real-time qRT-PCR tests (Appendix 1). Anonymized data, including age group, sex, municipality, month of serum sample collection, and test results, were collected. This study was approved by the Ethics Committee of the “Sindicato Médico y Ramas Afines de la Caja Nacional de Salud”, Cochabamba (No. 01/2024). Of the suspected cases, 5,923 (61.9%) tested positive by at least one method, representing 73.3% of all confirmed cases in the department. Cochabamba city reported the highest number of cases (4,059 out of 5,897 with available data; 68.8%). The outbreak peaked at the end of the rainy season, with 30.4% of total cases reported in April and 37.9% in May. The most affected age groups were 0–20 years (30.3%) and 21–40 years (37.3%) (Table 1).

**Table 1.** Characteristics of patients with suspected and confirmed dengue infection in Cochabamba, Bolivia.

| Variable | No. positive/<br>No. suspected cases (%) | No. positive/<br>No. confirmed cases (%) |
| --- | --- | --- |
| Result* |  |  |
| Positive | 5923/9576 (61.9) | NA |
| Negative | 3653/9576 (38.1) | NA |
| Sex |  |  |
| Female | 3314/9568 (34.6) | 3314/5920 (56.0) |
| Male | 2605/9568 (27.2) | 2605/5920 (44.0) |
| Municipality/Province |  |  |
| Cochabamba (2,558 m) | 4059/9422 (43.1) | 4059/5897 (68.8) |
| Quillacollo (2,422 m) | 294/9422 (3.1) | 294/5897 (5.0) |
| Capinota (2,386 m) | 236/9422 (2.5) | 236/5897 (4.0) |
| Sacaba (2,719 m) | 179/9422 (1.9) | 179/5897 (3.0) |
| Chimoré (240 m) | 153/9422 (1.6) | 153/5897 (2.6) |
| Mizque (2,007 m) | 151/9422 (1.6) | 151/5897 (2.6) |
| Month (year 2024) |  |  |
| January | 241/9576 (2.5) | 241/5923 (4.1) |
| February | 334/9576 (3.5) | 334/5923 (5.6) |
| March | 691/9576 (7.1) | 691/5923 (11.7) |
| April | 1803/9576 (18.6) | 1803/5923 (30.4) |
| May | 2243/9576 (23.2) | 2243/5923 (37.9) |
| June | 575/9576 (5.9) | 575/5923 (9.7) |
| July | 36/9576 (0.4) | 36/5923 (0.6) |
| Age-range (years) |  |  |
| 0-20 | 1784/9535 (18.7) | 1784/5902 (30.3) |
| 21-40 | 2204/9535 (23.1) | 2204/5902 (37.3) |
| 41-60 | 1288/9535 (13.5) | 1288/5902 (21.8) |
| >60 | 626/9535 (6.6) | 626/5902 (10.6) |

\*ELISA NS1, ELISA IgM or qRT-PCR tests
Abbreviations: NA, not applicable; m: meter above sea level

A total of 437 positive samples were previously serotyped at SEDES, with five identified as DENV-1 and 432 as DENV-2. These results align with data from the most recently reported epidemics in Bolivia, which have been driven by DENV-2 and DENV-1 (4). Historically, genotype III (Asian-American) was the only genotype of DENV-2 described in Bolivia, with evidence of sustained local circulation (11). However, the recent emergence of genotype II in several neighboring regions (12–14), along with DENV-2 genotype II sequences from Bolivia deposited in the GISAID database in 2023, suggests the introduction of this genotype into the country (Appendix 2). To determine the genotype of the virus lineages circulating in Cochabamba in 2024, we sequenced and analyzed seven nearly complete DENV-2 genomes (>85% of the coding DNA sequence [CDS]) from samples collected in June 2024 (Appendix 1). Six samples were collected from Cochabamba city, and one from Quillacollo (Appendix 3) (GenBank accession numbers PV426482 to PV426488). We reconstructed the phylogenetic relationships between virus genomes from Cochabamba in June 2024 and a set of reference sequences representative of DENV-2 genotypes, and found that all genomes belonged to genotype II (Appendix 4), similar to the Bolivian sequences from 2023 previously identified in GISAID.

To evaluate whether the 2024 Cochabamba outbreak was linked to DENV-2 lineages identified in Bolivia in 2023, we inferred a phylogeny based on a dataset combining all virus genomes from Cochabamba with all nearly complete genomes from DENV-2 genotype II available in GISAID as of February 23, 2025 (Appendix 2). The resulting phylogenetic tree (Figure 1), showed that the 2024 Cochabamba sequences formed two separate clades (A and B) with good statistical support (bootstrap >95), indicating that several, closely related, DENV-2-II lineages were responsible for the outbreak. Although both clades were phylogenetically close to other recent sequences from Bolivia sampled in Santa Cruz (2023), clade A was rooted by a sequence sampled in Parana state, Brazil, in 2024 (Curitiba, hDenV2/Brazil/SP-IAL-357154993/2023), and clade B by two other sequences from Brazil sampled in 2024 in the states of Paraná (hDenV2/Brazil/PR-Fiocruz-LRV24H3138/2024, Castavel) and Santa Catarina (hDenV2/Brazil/SC-LACENSC-422083052/2024, São Francisco do Sul), suggesting that this outbreak may have resulted from a new introduction into the country.

**Figure 1.**
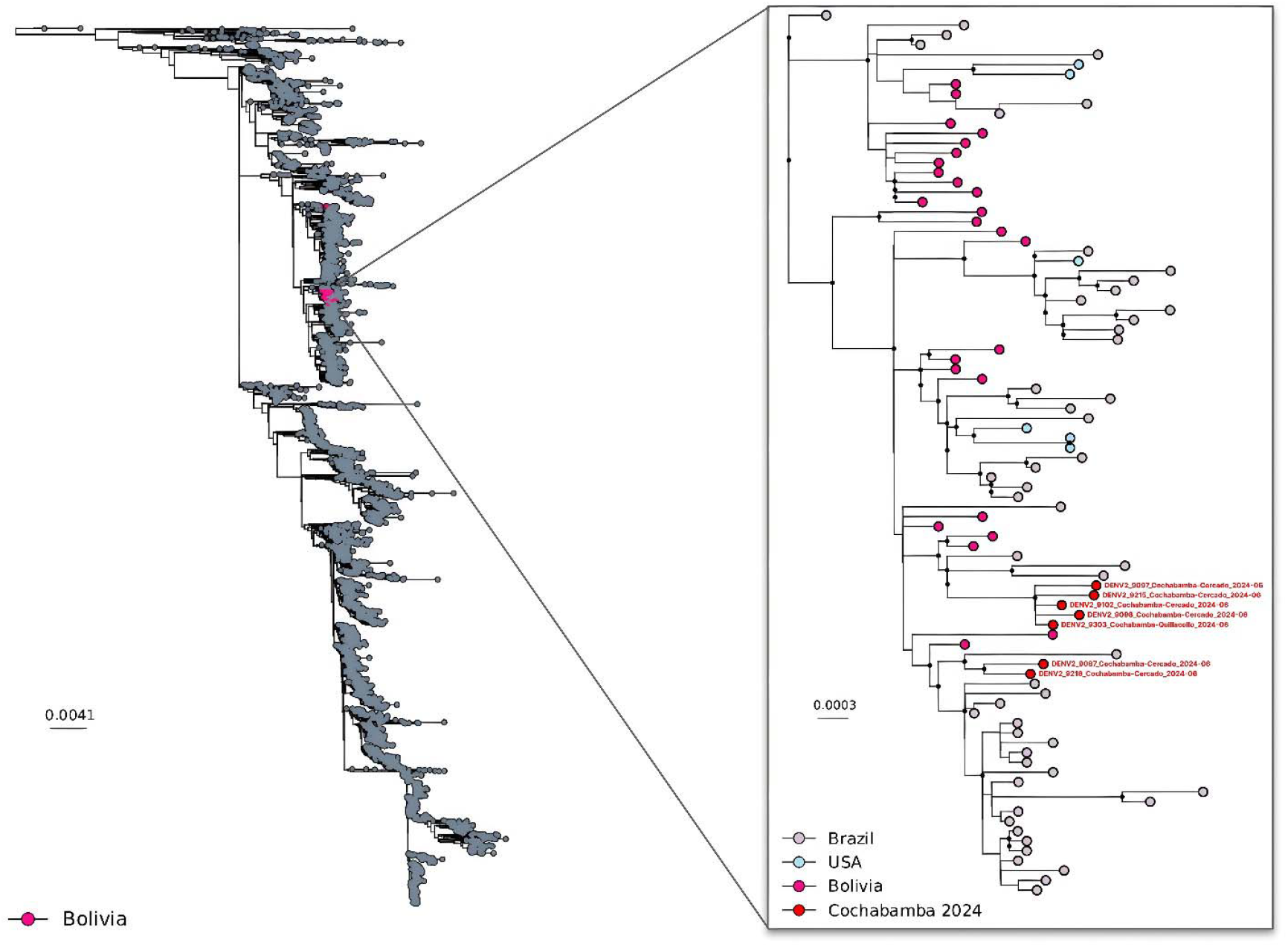
DENV-2-II phylogeny with a focus on sequences from Cochabamba, 2024. Maximum-Likelihood phylogeny of all genomes from Cochabamba 2024 combined with all DENV-2 sequences from genotype II covering more than 85 % of the CDS available on GISAID as of the 23rd of February 2025. Phylogenetic inference was performed using IQTREE2 using a GTR+F+R5 model (General time reversible model with empirical base frequencies and a FreeRate model with 5 categories) with ultrafast bootstrap approximation (1000 replicates). The tree was rooted using the divergent strain QML22 (KX274130). In the subtree in the right-hand panel, nodes with a bootstrap support above 95 are highlighted with a black circle.

## Conclusions

This study reports a significant rise in dengue cases in Cochabamba, Bolivia, in 2024, highlighting the growing challenge of controlling the disease as it spreads into previously unaffected areas. This surge is closely linked to the geographic expansion of *Ae. aegypti*, likely driven by climate change, which has altered environmental conditions in ways that enhance mosquito survival and breeding. The establishment of *Ae. aegypti* at high altitudes has enabled DENV transmission in populations with historically low levels of immunity (15).

Our findings provide valuable insights into the current viral lineages circulating in Bolivia, confirming the local emergence of genotype II, consistent with patterns observed in several other countries in the region (12–14). Phylogenetic analysis indicates that the 2024 Cochabamba outbreak was associated with at least two separate virus lineages, distinct from identified in the country in 2023. This suggests that DENV circulation in Cochabamba stemmed from multiple recent introduction events. Clades A and B closest phylogenetic relatives were found in Brazil, suggesting these clades were seeded from neighboring countries. However, due to the limited availability of recent DENV genomic data for Bolivia and several neighboring countries, including Paraguay and Argentina, the precise source of these introductions remains uncertain.

The exceptional circumstances of the Cochabamba outbreak, with substantial virus transmission occurring at high altitudes and lower temperatures, raise the question of a potential viral adaptation to less favorable environmental conditions. Further genomic analyses, combined with the experimental characterization of virus strains described in this study, are warranted to evaluate whether viral adaptation contributed to the increased DENV circulation observed in Cochabamba in 2024.

This study emphasizes the need to strengthen and adapt dengue control and surveillance strategies to align with the virus’s evolution, expanding range, and shifting transmission dynamics. A multifaceted approach is essential, including effective vaccines, sustainable vector control strategies, public health education, and coordinated monitoring of viral diversity at regional and international levels.

## Data Availability

All data produced in the present work are contained in the manuscript

## Acknowledgments

We would like to thank the staff of the Servicio Departamental de Salud (SEDES) in Cochabamba, Bolivia, for their assistance with sample collection. We also thank Bernard Tenebray, Thomas Canivez, Laurent Bosio, Manon Geulen, Manon Peden, Melissa Rupert, and Melissa Venot from the Unité des Virus Émergents and the National Reference Center for Arboviruses for their technical assistance, as well as Elizabeth Villarroel Romero from the Central Laboratory in Cochabamba, Bolivia.

## Biographical Sketch

Dr. Saba Villarroel is a postdoctoral researcher at Mahidol University in Thailand. Her primary research interests include the diagnosis and epidemiology of infectious diseases, with a particular focus on emerging and re-emerging viruses.

