## Appendix for "Dengue virus outbreak in Cochabamba, Bolivia"

### Appendix 1. Methods

*Serum collection and dengue testing*

Blood samples were collected via venipuncture at the National Health Service (SEDES) after obtaining signed informed consent, using serum collection tubes. Following centrifugation, the sera were analyzed for dengue virus according to the duration of the patient's symptoms. For individuals presenting within five days of symptom onset, detection was performed using either RT-qPCR with CDC primers (DENV-1–4 Real-Time rRT-PCR Multiplex Assay) or the commercial NS1 antigen ELISA test (Euroimmun, Lübeck, Germany). For patients with symptoms lasting more than six days, an in-house MAC-ELISA was used.

*Virus sequencing*

A specific set of primers was used to generate eight overlapping amplicons spanning the entire DENV-2 genome with the Superscript IV one step RT-PCR System (ThermoFisher Scientific). PCR mixes (final volume 25µL) contained 3µL of nucleic acid extract, 1,25µL of each primer (10µM), 12.5 µL of 2X Platinum SuperFi RT-PCR Master Mix, 6.5µL of RNAse free water and 0.5µL of SuperScript IV RT Mix. Amplifications were performed using the following conditions: 10 min at 55°C, 2 min at 98°C, followed by 40 cycles with the three following steps: 10 sec at 98°C, 10 sec at 55°C and 1.45 min at 68°C, and a final step at 68°C for 5 min.

The size of PCR products was controlled by gel electrophoresis. For each sample, an equimolar pool of all amplicons was prepared and purified using Monarch PCR & DNA Cleanup Kit (New

England Biolabs). After Qubit quantification using the Qubit® dsDNA HS Assay Kit and Qubit 2.0 fluorometer (Thermo Fisher) amplicons were sonicated (Bioruptor®, Diagenode, Liège, Belgium) into 250pb long fragments. Fragmented DNA was used for library building using the Ion Plus Fragment Library Kit with the AB Library Builder System (Thermo Fisher). To ensure the equimolar pooling of the barcoded samples, a real-time PCR quantification step was performed using Ion Library TaqMan™ Quantitation Kit (Thermo Fisher). An emulsion PCR of the pools was performed, followed by loading on 530 chips using the automated Ion Chef instrument (Thermo Fisher), and sequencing using the S5 Ion torrent technology (Thermo Fisher), following manufacturer's instructions.

Read data were analyzed with an in-house Snakemake pipeline (1). Read alignment was achieved using BWA MEM (v0.7.17 (2)) using, as a reference, the best match identified by blasting (magicblast, v1.7.7 (3)) sequencing reads using a database of flavivirus sequences including 42 sequences representative of DENV-2 genetic diversity. Consensus sequences were called using the ivar (v1.3.1 (4)) consensus command, and a minimum coverage depth of 50x. Regions with insufficient coverage were masked with N characters.

*Virus sequence data analysis*

First, we aligned our DENV-2 genomes and a set of reference sequences representative of DENV-2 genotypes and inferred their phylogenetic relationships with IQ-Tree (version 1.6.12 (5,6), using the best-fit model identified by ModelFinder and assessed branch support using an ultrafast bootstrap approximation (UFBoot2) (1000 replicates). We found that they all belonged to DENV-2 genotype II (also called Cosmopolitan). The set of reference genomes used for genotyping was selected based on the set of genomes used in the widely-used tool genome detective to DENV genotyping (7). For the set of reference genomes to better reflect the phylogenetic diversity within DENV-2 serotype we added the sequence of the highly divergent strain QML22 isolated from a traveler returning from Borneo to Australia (8).

All publicly available sequences for DENV-2 genotype II with a length above 8500 nt were downloaded from the GISAID database on February, 23rd, 2025. The sequences were aligned using MAFFT (version 7.51142), trimmed to their coding regions (ORF) and inspected manually. We then removed potential recombinant sequences from the dataset using the Recombination Detection Program (RDP) version 4. We used RDP, GENCONV and MAXCHI methods for primary screening and BOOTSCAN and SISCAN methods to check for recombination signals (9–13). We used the automask option to ensure optimal recombination detection.

Using the resulting recombinant-free alignments, we performed ML phylogenetic reconstruction with IQ-Tree (version 1.6.1250), using a GTR+F+R4 model (General time reversible model with empirical base frequencies and a FreeRate model with 5 categories) and assessed branch support using an ultrafast bootstrap approximation (UFBoot2) (1000 replicates).

**Appendix 2. Data Availability and data snapshot**

GISAID Identifier: EPI_SET_250223hf; doi: 10.55876/gis8.250223hf

All genome sequences and associated metadata in this dataset are published in GISAID's EpiArbo database. To view the contributors of each individual sequence with details such as accession number, Virus name, Collection date, Originating Lab and Submitting Lab and the list of Authors, visit 10.55876/gis8.250223hf

EPI_SET_250223hf is composed of 7,409 individual genome sequences.

The collection dates range from 1982-01-01 to 2025-01-20;

Data were collected in 52 countries and territories;

All sequences in this dataset are compared relative to the official reference sequence employed by GISAID.

**Appendix 3. Sequence data and corresponding metadata for the seven sequenced samples from Cochabamba, Bolivia**

| Sample number | Virus | Sampling year-month | Country of origin | Geographical details Department (Province) | GenBank Accession number | %genome covered | Genotype | Lineage |
|---|---|---|---|---|---|---|---|---|
| 9303 | DENV-2 | 2024-06 | Bolivia | Cochabamba (Quillacollo) | PV426488 | 99.38 | II (Cosmopolitan) | F.1.2 |
| 9102 | DENV-2 | 2024-06 | Bolivia | Cochabamba (Cercado) | PV426487 | 99.43 | II (Cosmopolitan) | F.1.2 |
| 9098 | DENV-2 | 2024-06 | Bolivia | Cochabamba (Cercado) | PV426486 | 99.4 | II (Cosmopolitan) | F.1.2 |
| 9097 | DENV-2 | 2024-06 | Bolivia | Cochabamba (Cercado) | PV426485 | 99.43 | II (Cosmopolitan) | F.1.2 |
| 9087 | DENV-2 | 2024-06 | Bolivia | Cochabamba (Cercado) | PV426484 | 99.43 | II (Cosmopolitan) | F.1.2 |
| 9215 | DENV-2 | 2024-06 | Bolivia | Cochabamba (Cercado) | PV426482 | 84.83 | II (Cosmopolitan) | F.1.2 |
| 9218 | DENV-2 | 2024-06 | Bolivia | Cochabamba (Cercado) | PV426483 | 86.75 | II (Cosmopolitan) | F.1.2 |

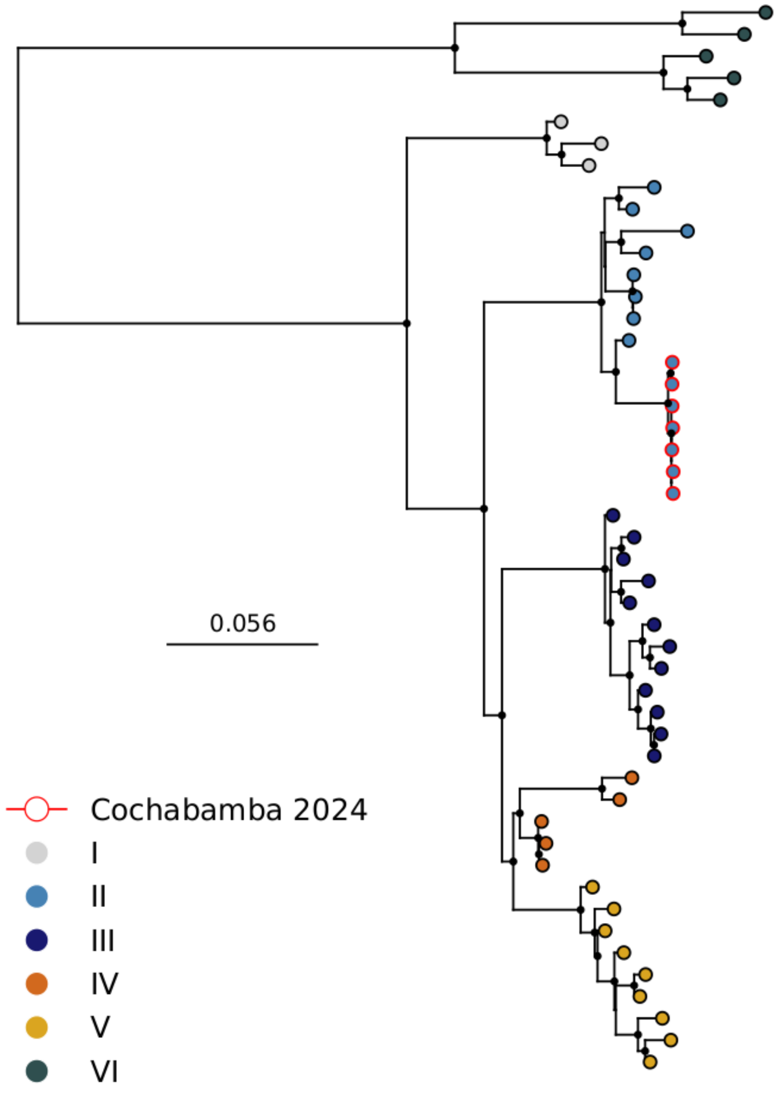


**Appendix 4.** Maximum-Likelihood phylogeny of all DENV-2 sequences from the Cochabamba 2024 epidemic with a set of reference sequences representative of DENV-2 genotypes Phylogenetic inference was performed using IQTREE2 under the best substitution model identified by ModelFinder with ultrafast bootstrap approximation (1000 replicates). The tree was rooted using the highly divergent strain QML22 (KX274130). Nodes with bootstrap support above 95 are highlighted with a black circle.
